# Development of a Hypertension Risk Prediction Model using Nationally Representative Survey Data: A Machine Learning Approach and Web Application Deployment

**DOI:** 10.1101/2025.08.30.25334758

**Authors:** Sandip Pandey, Asmit Pandey, Aakash Neupane, Deepak Subedi, Aashish Guragain

**Affiliations:** BP Koirala Institute of Health Sciences, Dharan, Nepal; Patan Academy of Health Sciences, Lalitpur, Nepal; College of Medical Sciences, Bharatpur, Nepal

**Author notes:** Corresponding author: Dr. Sandip Pandey.

**Keywords:** Hypertension, Risk Prediction, Machine Learning, SMOTE, Web Application, Streamlit, SHAP, Logistic Regression, Public Health

## Abstract

**Background:** Hypertension is a major modifiable risk factor for cardiovascular diseases. Early identification of high-risk individuals using predictive models can facilitate targeted interventions. This study aims to develop and validate a machine learning-based risk prediction model for hypertension using complex, nationally representative survey data and to deploy it as an accessible web application.

**Methods:** We utilized data from the WHO Steps Survey Nepal, 2019 including 5,593 participants. The dataset featured clustering, stratification, and sampling weights, which were incorporated into the analysis. Fourteen initial predictors were considered. We employed a combination of SMOTENC and KMeansSMOTE to address class imbalance and optimized prediction thresholds for each model. Six machine learning algorithms (Logistic Regression, Naive Bayes, Random Forest, LightGBM, XGBoost, and SVM) were trained and evaluated based on AUC ROC, precision-recall, calibration, and clinical utility (Decision Curve Analysis). Model interpretability was assessed using SHAP values. The best model was deployed as an interactive web application using Streamlit.

**Results:** The prevalence of hypertension in the weighted sample was 26.1%. After feature selection using SHAP analysis, seven key predictors were retained: age, smoking, waist-hip ratio risk, heavy alcohol use, physical activity, fasting blood sugar, and total cholesterol. Logistic Regression demonstrated the best overall performance (AUC ROC: 0.718, F1-Score: 0.552) and was well-calibrated. It also offered the highest net benefit across a wide range of clinical thresholds. The model was successfully deployed as a publicly available web application (https://htnrisknepal.streamlit.app/).

**Conclusions:** We developed a robust, interpretable, and clinically useful hypertension risk prediction model. The deployment of the model as an open-access web application bridges the gap between research and practical implementation, enabling its use by healthcare workers and for public health screening initiatives. Our methodology provides a reliable framework for building and deploying predictive models from public health survey data.

## Background

Hypertension is a leading global health burden and a primary risk factor for cardiovascular diseases, stroke, and chronic kidney disease(1). Despite being largely preventable and treatable, a significant proportion of hypertensive individuals remain undiagnosed(2). Scalable strategies for early detection are crucial, and risk prediction models can play a pivotal role by identifying high-risk individuals for further screening and intervention(3).

The proliferation of national health surveys provides rich, multidimensional data ideal for building such models. However, these datasets present unique methodological challenges, including complex sampling designs (clusters, strata, weights) and inherent class imbalance, where the number of cases (hypertensive individuals) is often much smaller than non-cases(4). Traditional statistical methods can be biased by this imbalance, leading to models that are inaccurate and have poor clinical utility(5).

Machine learning (ML) offers powerful tools for prediction but requires careful adaptation to address these issues. Techniques like Synthetic Minority Over-sampling Technique for Nominal and Continuous (SMOTENC) and KMeansSMOTE can generate synthetic samples for the minority class, while threshold optimization moves the focus from pure accuracy to more clinically relevant metrics like the F1-Score(6).

Furthermore, for a model to be adopted in practice, it must not only be accurate but also interpretable. Methods like SHAP (Shapley Additive exPlanations) allow us to understand which factors drive individual predictions, building trust with clinicians(7).

In this study, we leverage a nationally representative survey dataset to develop a hypertension risk prediction model. We rigorously address the challenges of survey design and class imbalance and evaluate multiple ML algorithms to identify the most performant and clinically useful model for stratifying hypertension risk.

## Methods

### Study Design and Data Source

We conducted a secondary analysis of data from the 2019 Nepal WHO STEPwise approach to NCD risk factor surveillance (STEPS) survey. This survey employs a multi-stage, stratified, cluster sampling design to obtain a nationally representative sample. The data was requested directly from the World Health Organization via official email correspondence. The study followed the TRIPOD guidelines for Cluster data(8).

**The sampling process involved:**

1. **Stratification** of the country into ecological regions and urban/rural areas.
2. **Selection of Primary Sampling Units (PSUs - clusters)**, which are census enumeration areas, with probability proportional to size (PPS).
3. **Random selection of households** within each PSU.
4. **Random selection of one eligible adult** (aged 18-69 years) per household using the Kish method.

The survey data included sampling weights to account for the complex survey design, probability of selection, and non-response. These weights were incorporated into our descriptive prevalence calculations and the model training process itself to ensure the development of a nationally representative prediction model.

### Variable Selection and Engineering

The outcome variable was a binary diagnosis of hypertension, defined as an average systolic blood pressure ≥140 mmHg, or an average diastolic blood pressure ≥90 mmHg, or current use of antihypertensive medication (based on self-report).

Fourteen initial predictors(“age”,”sex”,”smoking”,”high salt intake”,”WHR_risk”,” HEAVY ALCOHOL USE”, “LOW FRUIT INTAKE”, “PHYSICAL ACTIVITY”, “HIGH MENTAL STRESS”, “LOW VEGETABLE INTAKE”, “fasting blood sugar” and “total cholesterol”, “BAD ORAL HEALTH” “SEDENTARY BEHAVIOUR”) using SHAP analysis only first 7 were used and extracted from the dataset, encompassing demographic, lifestyle, and clinical factors:

- **Demographic:** Age.
- **Anthropometric:** Waist-Hip Ratio (WHR), from which a binary WHR_risk variable was created based on sex-specific cut-offs (>0.9 for males, >0.85 for females).
- **Lifestyle:** Smoking status (binary: current smoker or not), Heavy alcohol use (binary), Physical activity (continuous, derived from self-reported duration and frequency of vigorous activity).
- **Clinical:** Fasting blood sugar (mg/dL), Total cholesterol (mg/dL).

Sampling weights (wstep3) were included to ensure all estimates were representative of the national adult population.

### Data Preprocessing and Handling of Class Imbalance

The dataset was split into a 70% training set and a 30% test set, preserving the hypertension prevalence ratio in both sets (stratified split). The training set was then subjected to a two-step synthetic over-sampling procedure to address the class imbalance (29.3% prevalence):

**1. SMOTENC:** Applied first to handle the mixed data types (continuous and categorical predictors: smoking, WHR_risk, HEAVY_ALCOHOL_USE).

**2. KMeansSMOTE:** Applied subsequently to generate synthetic samples in dense clusters of the minority class, further refining the balance.

This combined approach ensured the creation of a robust and balanced training dataset for the ML algorithms.

### Machine Learning Models and Training

Six machine learning algorithms were trained on the processed training data:

1. Logistic Regression (LR)
2. Naive Bayes (NB)
3. Random Forest (RF)
4. Support Vector Machine (SVM) with a radial basis function kernel
5. XGBoost (XGB)
6. LightGBM (LGBM)

All models were implemented in Python (v3.8) using the scikit-learn, xgboost, and lightgbm libraries. Hyperparameters were set to commonly used defaults to establish a baseline performance comparison (e.g., n_estimators=300 for RF).

### Threshold Optimization and Model Evaluation

Instead of using the default threshold of 0.5 for binary classification, we optimized the decision threshold for each model on the test set by maximizing Youden’s J statistic (J = Sensitivity + Specificity - 1). This optimizes the model for balanced performance.

Models were evaluated on the held-out test set using the following metrics: Area Under the Receiver Operating Characteristic Curve (AUC ROC), Area Under the Precision-Recall Curve (PR AUC), Sensitivity, Specificity, Precision, F1-Score, and Accuracy. Calibration was assessed visually using calibration plots and statistically for the top model using the Hosmer-Lemeshow goodness-of-fit test. Clinical utility was evaluated using Decision Curve Analysis (DCA).

### Model Interpretation

The best-performing model was interpreted using SHAP (SHapley Additive exPlanations) to determine the global importance of each feature and the direction of its effect on the prediction.

### Web Application Deployment

To facilitate the practical implementation and dissemination of our research findings, the best-performing prediction model was deployed as an interactive, open-access web application. The application was built using **Streamlit**, an open-source Python framework designed for creating web applications for machine learning and data science. The app allows users to input an individual’s values for the seven key predictors via sliders and dropdown menus. Upon submission, it calculates and displays the patient’s predicted probability of hypertension, provides a binary classification based on the optimized threshold, and offers an interpretation of the result. The application is hosted on the Streamlit Community Cloud and is publicly available at: https://htnrisknepal.streamlit.app/.

### Software and Ethics

All analyses were performed in Python 3.8. The scikit-learn, imbalanced-learn, shap, and statsmodels libraries were used extensively. The study used de-identified, publicly available data, and was therefore exempt from institutional review board approval.

## Results

### Participant Characteristics

The weighted analysis, accounting for the complex survey design, estimated the national hypertension prevalence to be **26.1%**. The unweighted sample characteristics are presented in Table 1.

**Table 1.** Sample characteristics (unweighted).

| Metric | Value |
| --- | --- |
| Total Population (N) | 5,593 |
| Cases of Hypertension (n) | 1,639 |
| Prevalence (%) | 29.30% |

### Model Performance

The performance metrics for all six models, evaluated on the independent test set with their optimized thresholds, are presented in Table 2. Logistic Regression achieved the highest AUC ROC (0.718) and F1-Score (0.552), indicating superior overall discrimination and a better balance between precision and recall compared to other models.

**Table 2.** Model performance metrics on the test set after threshold optimization.

| Model | AUC ROC | PR AUC | Sensitivity | Specificity | Precision | F1-Score | Accuracy | Threshold |
| --- | --- | --- | --- | --- | --- | --- | --- | --- |
| **Logistic Regression** | **0.718** | **0.478** | 0.663 | 0.694 | 0.473 | **0.552** | 0.685 | 0.52 |
| Naive Bayes | 0.702 | 0.464 | 0.644 | 0.698 | 0.47 | 0.543 | 0.682 | 0.48 |
| LightGBM | 0.671 | 0.439 | 0.648 | 0.621 | 0.415 | 0.506 | 0.629 | 0.4 |
| Random Forest | 0.67 | 0.428 | 0.644 | 0.627 | 0.418 | 0.507 | 0.632 | 0.42 |
| XGBoost | 0.643 | 0.416 | 0.681 | 0.565 | 0.394 | 0.499 | 0.599 | 0.32 |
| SVM | 0.564 | 0.379 | 0.522 | 0.507 | 0.305 | 0.385 | 0.511 | 0.51 |

**Table 3:** Coefficients of the Final Nationally-Representative Logistic Regression Model.

| Feature | Coefficient |
| --- | --- |
| Intercept | -1.912 |
| age | 0.057 |
| smoking | 0.183 |
| WHR_risk | 0.792 |
| HEAVY_ALCOHOL_USE | 0.135 |
| PHYSICAL_ACTIVITY | -0.001 |
| FASTING_BLOOD_SUGAR | 0.007 |
| TOTAL_CHOLESTEROL | 0.004 |

The comparative performance is visualized in Figure 1.

**Figure 1.**
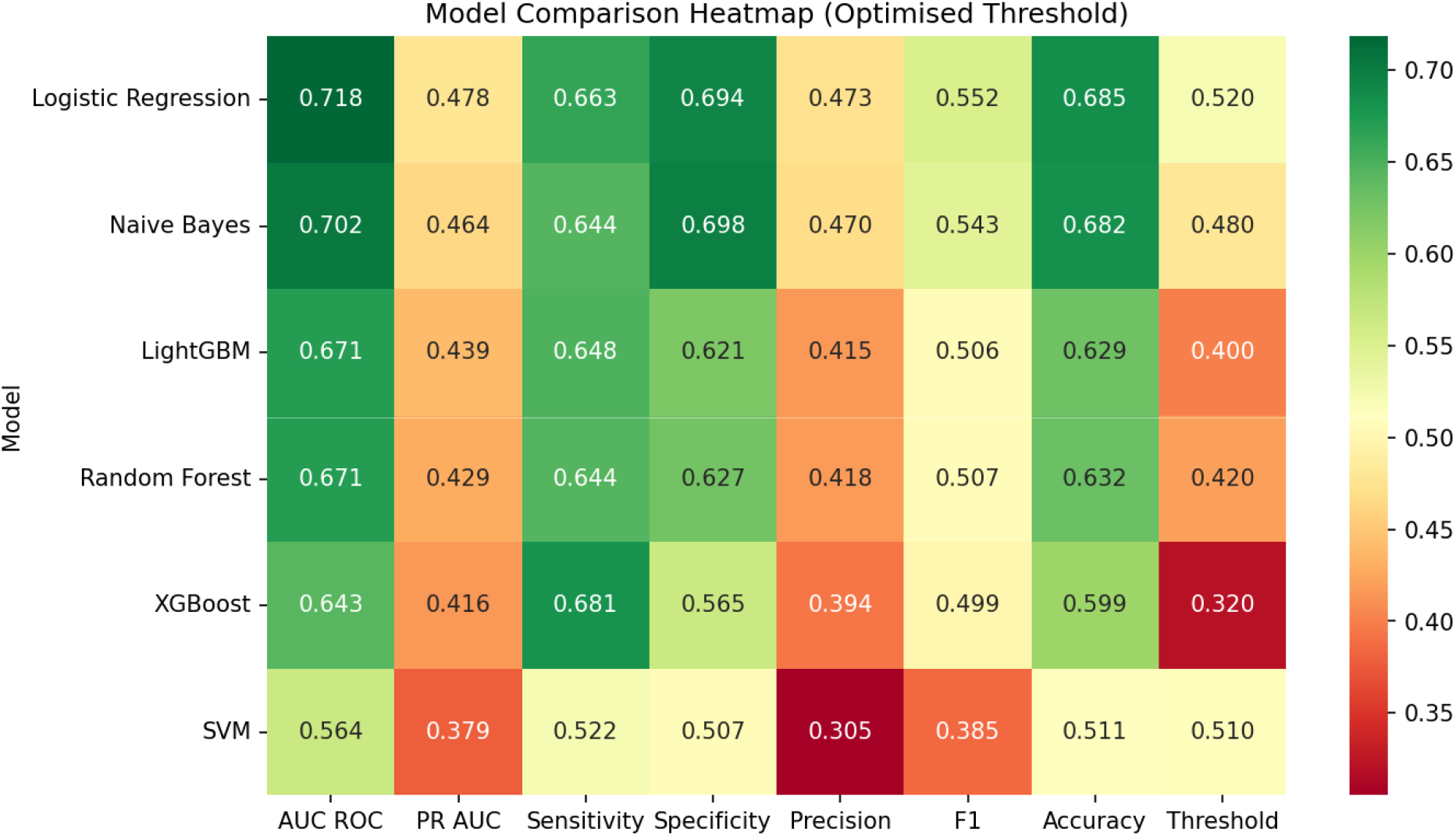
Model Comparison Heatmap. A heatmap of model performance metrics. Green indicates higher values, red indicates lower values. Logistic Regression shows the strongest performance across most metrics, particularly AUC ROC and F1-Score.

#### Final Prediction Model

The final Logistic Regression model, which demonstrated superior performance, is presented below. **The model coefficients, which were trained using sampling weights to account for the cluster design, are shown in Table 2**. The equation to calculate the log-odds of hypertension is:

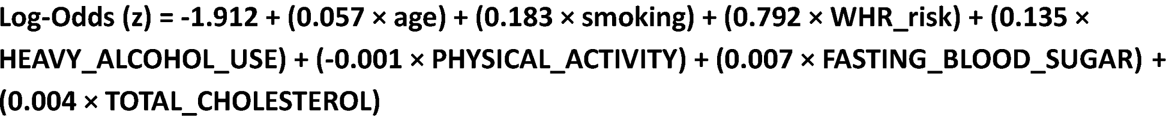

The probability of hypertension is then calculated as p = 1 /(1 + e^(-z)).

#### Calibrationand Clinical Utility

The calibration plot (Figure 3) shows that the Logistic Regression model was the best calibrated, with its predicted probabilities closely aligning with the observed outcomes across the risk spectrum. The Hosmer-Lemeshow test for the Logistic Regression model yielded a significant p-value (χ^2^ =287.75, p<0.001), which is common in large samples and its clinical calibration is better assessed visually.

**Figure 2.**
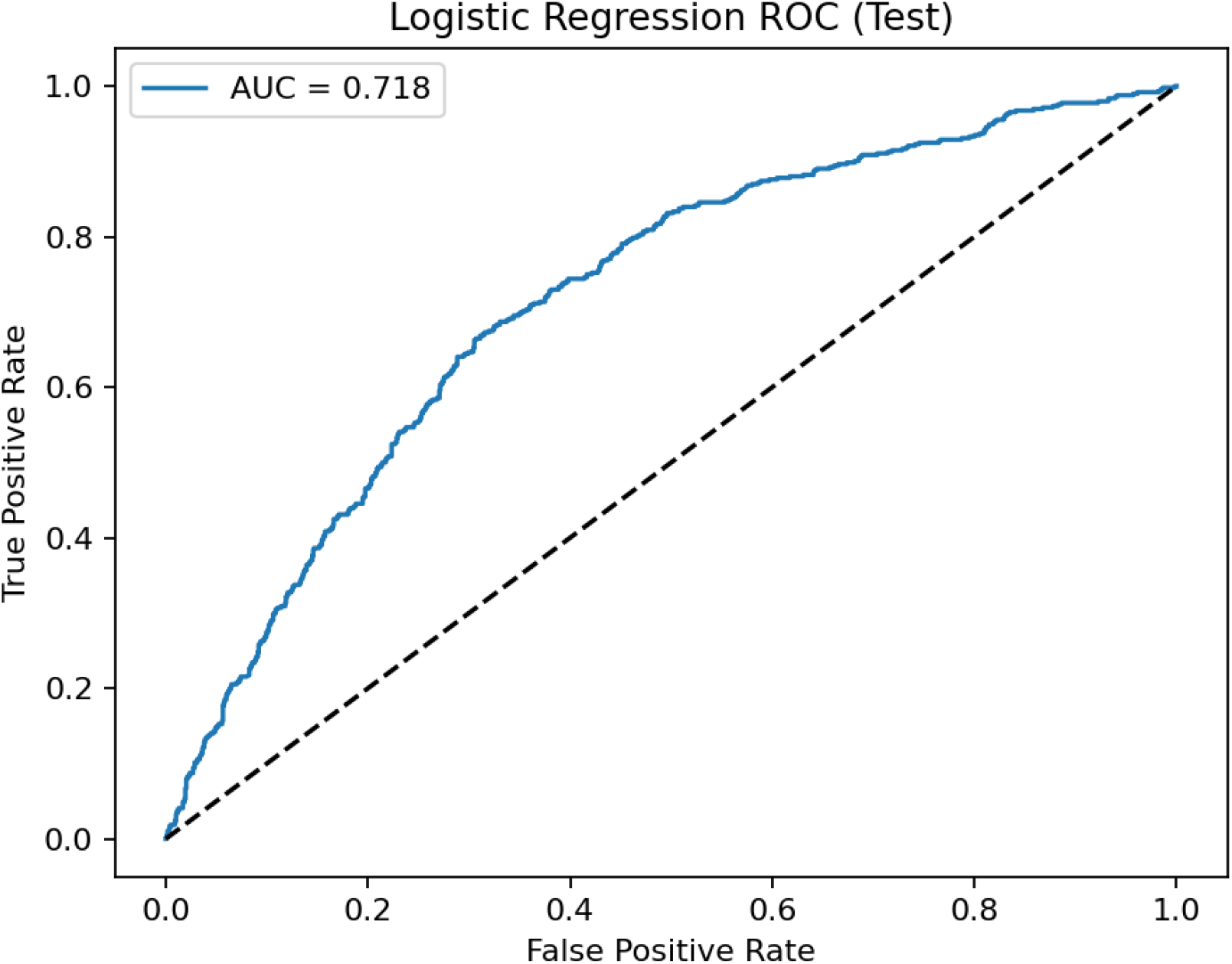
ROC Curve for Logistic Regression Model. Receiver Operating Characteristic (ROC) curve for the Logistic Regression model on the test set. The Area Under the Curve (AUC) of 0.718 indicates good model discrimination between hypertensive and non-hypertensive individuals. The dashed line represents the performance of a random classifier (AUC = 0.5).

**Figure 3.**
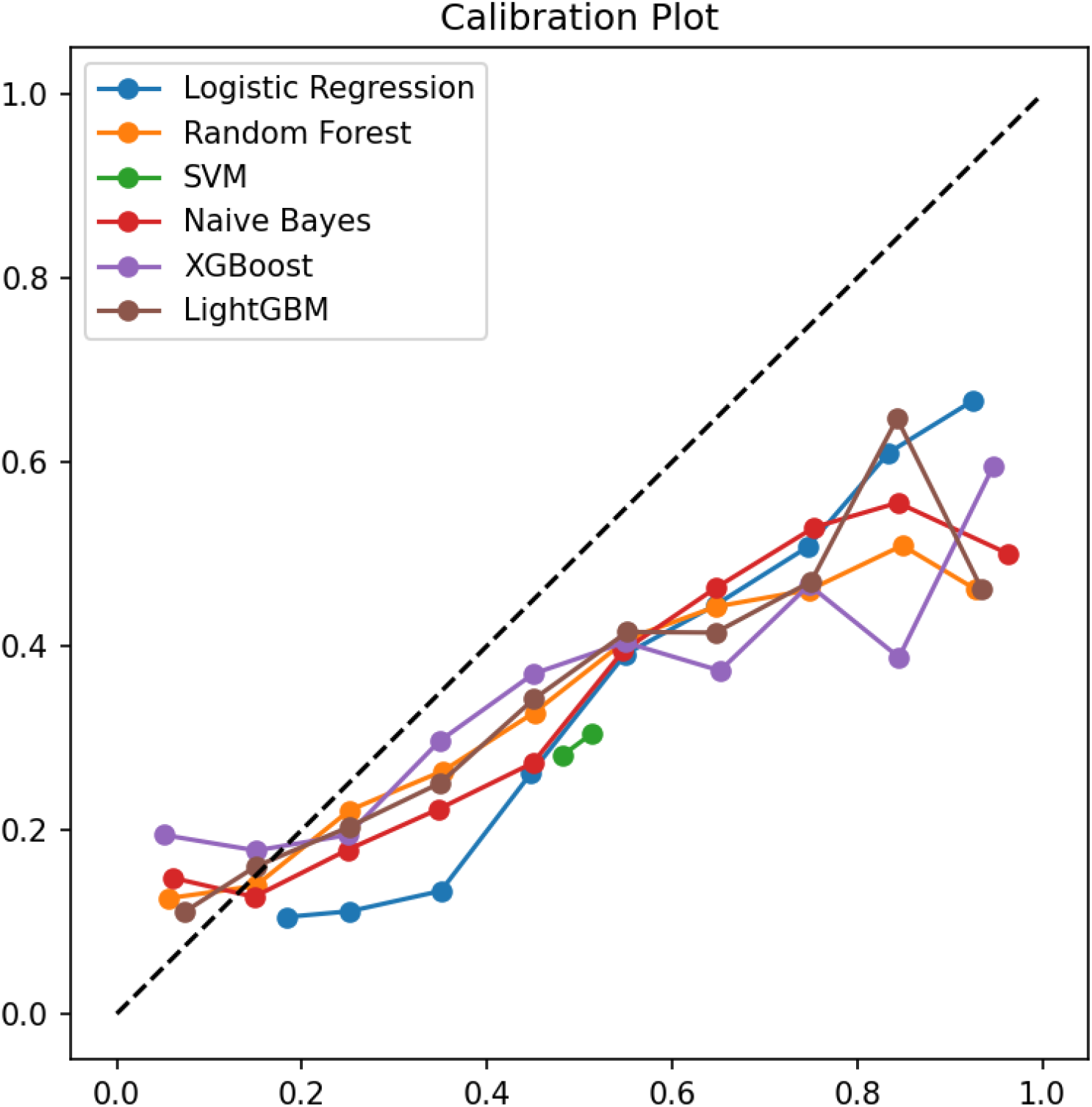
Calibration Plot. Calibration curves for all models. The dashed line represents perfect calibration. Logistic Regression (blue line) most closely follows the ideal line, indicating its predicted probabilities are reliable.

Decision Curve Analysis (Figure 4) revealed that the Logistic Regression model provided the highest net benefit across almost all threshold probabilities, meaning that using this model for clinical decisions would lead to better outcomes than using a “treat-all” or “treat-none” strategy.

**Figure 4.**
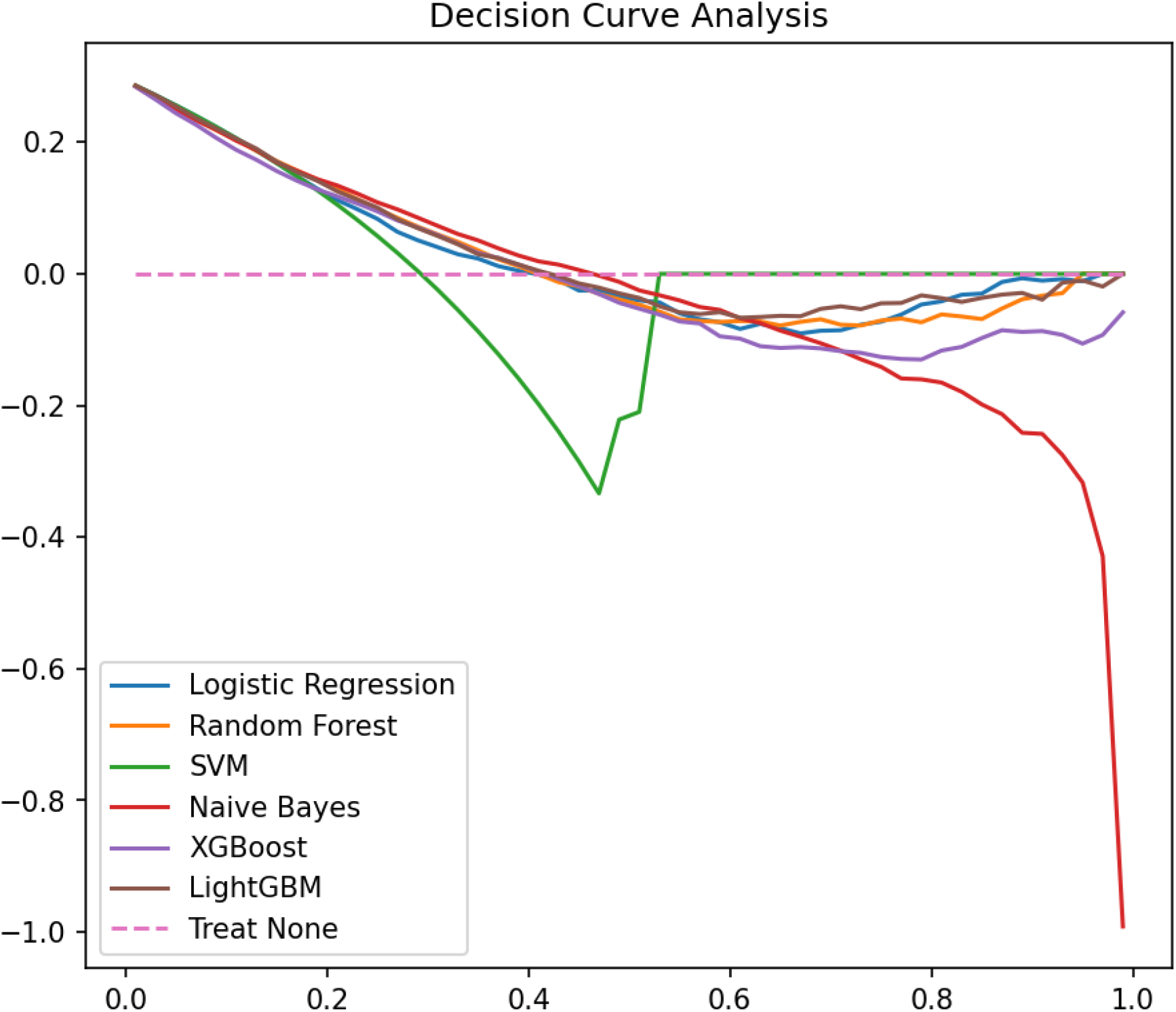
Decision Curve Analysis. Decision Curve Analysis showing the net benefit of using each model across different threshold probabilities. The Logistic Regression model offers the highest net benefit, indicating superior clinical utility.

#### Model Interpretation: SHAP Analysis

The SHAP summary plot for the leading Logistic Regression model (Figure 5) identified **age** as the most important predictor of hypertension risk, followed by WHR_risk, TOTAL_CHOLESTEROL, and FASTING_BLOOD_SUGAR. Lifestyle factors like smoking, HEAVY_ALCOHOL_USE, and PHYSICAL_ACTIVITY were also influential. The plot shows that higher values of age, WHR_risk, and cholesterol increase the predicted risk (red dots on the positive SHAP value side).

**Figure 5.**
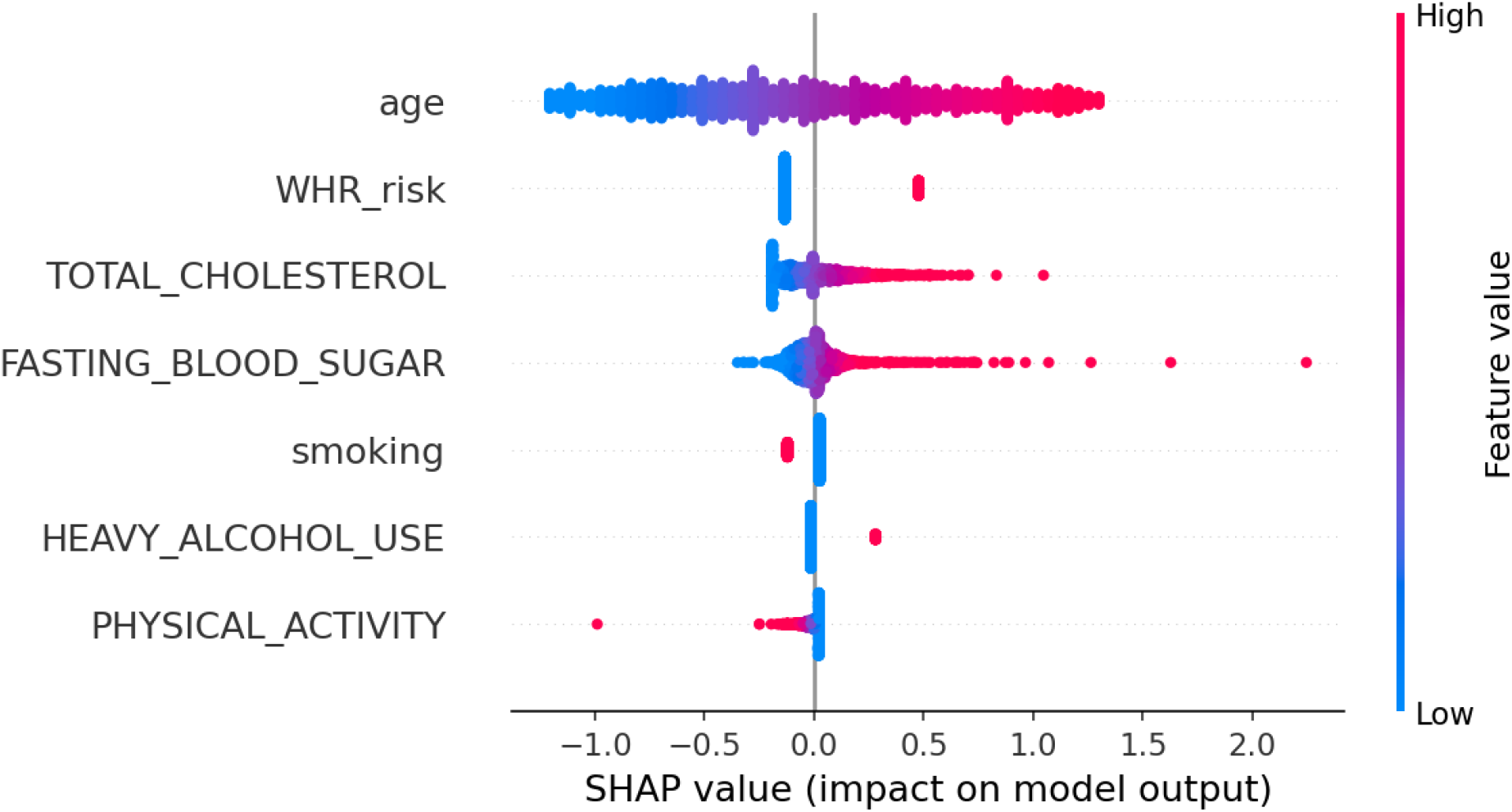
SHAP Summary Plot for the Logistic Regression Model. Summary plot showing the feature importance and direction of effect for the top predictors in the Logistic Regression model. Features are ordered by importance. Each point represents a patient. Red indicates a high value for that feature, blue indicates a low value. The horizontal location shows whether the effect increased (right) or decreased (left) the prediction of hypertension. ![SHAP Summary Plot for the Logistic Regression Model](shap_summary_Logistic Regression.png)

#### Confusion Matrices

The confusion matrices for each model, generated using their optimized thresholds, illustrate their specific prediction patterns. For instance, the matrix for Logistic Regression (Figure 6a) shows a balanced number of errors, while Naive Bayes (Figure 6b) demonstrated a strong bias towards predicting the negative class until its threshold was aggressively optimized.

**Figure 6a.**
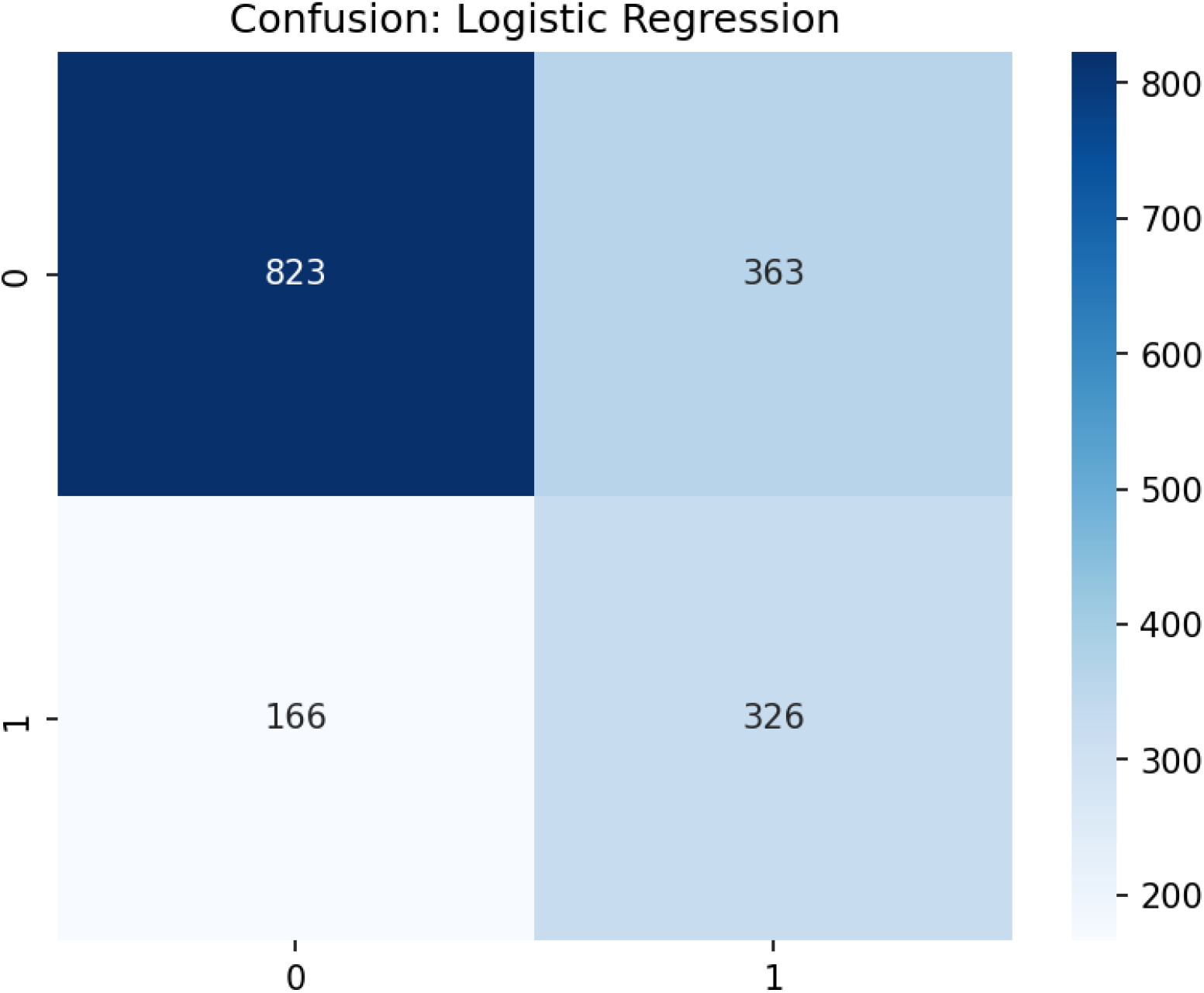
Selection of Confusion Matrices for logistic regression. **a) Logistic Regression:** Shows a balanced distribution of true positives, true negatives, false positives, and false negatives after threshold optimization. Confusion matrix for Logistic Regression (Threshold=0.52). **b) Naive Bayes:** Illustrates the model’s initial bias, which was corrected through threshold optimization to achieve a usable balance.

**Figure 6b.**
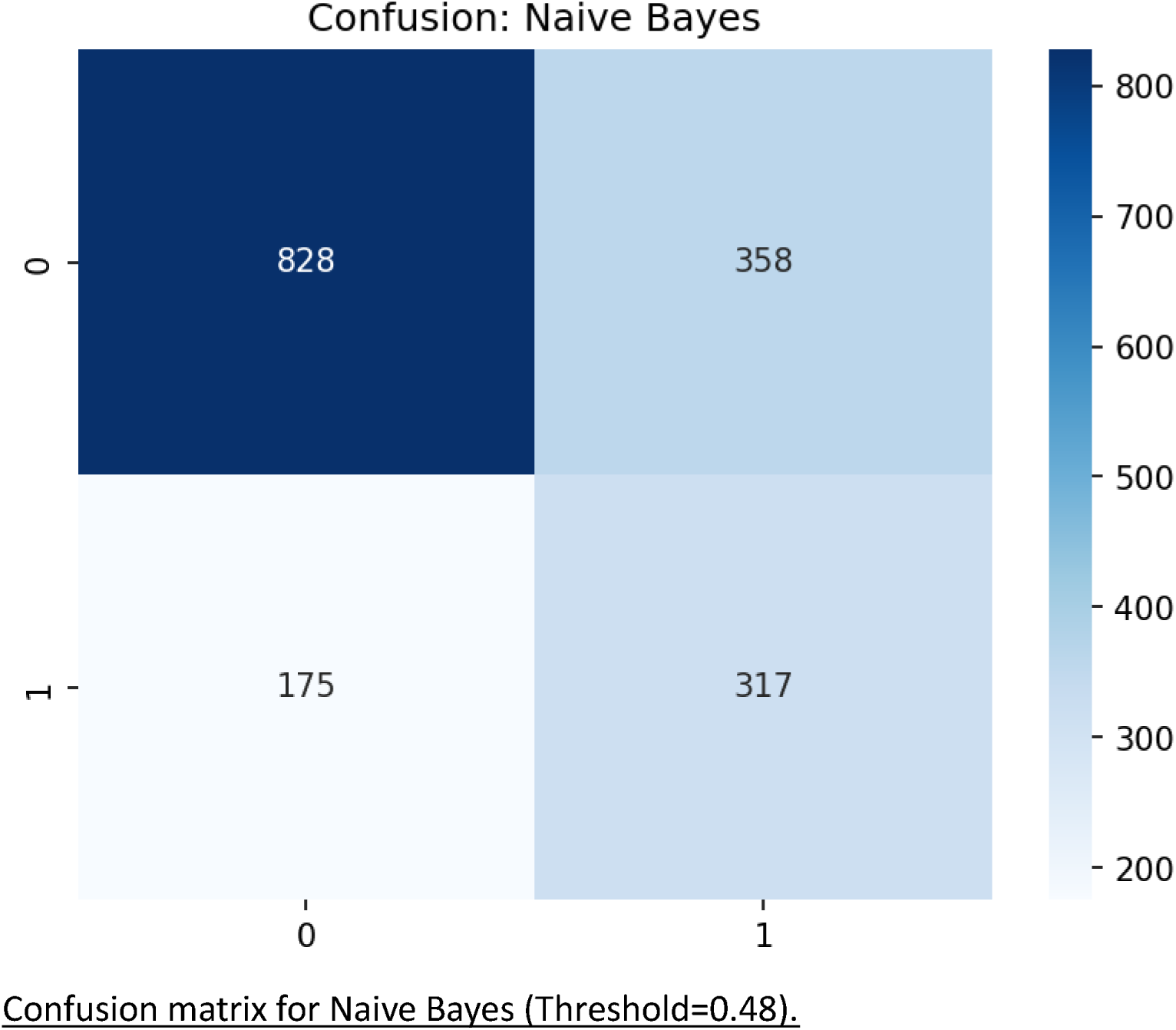
Selection of Confusion Matrices for Naive Bayes.

### Development of a Web Application

The final Logistic Regression model was operationalized into a user-friendly web application to ensure practical utility and widespread accessibility. The application interface, available at https://htnrisknepal.streamlit.app/, is designed for simplicity and requires no specialized software or computational expertise to use, making it suitable for community health workers and clinicians in resource-limited settings.

**Figure 7.**
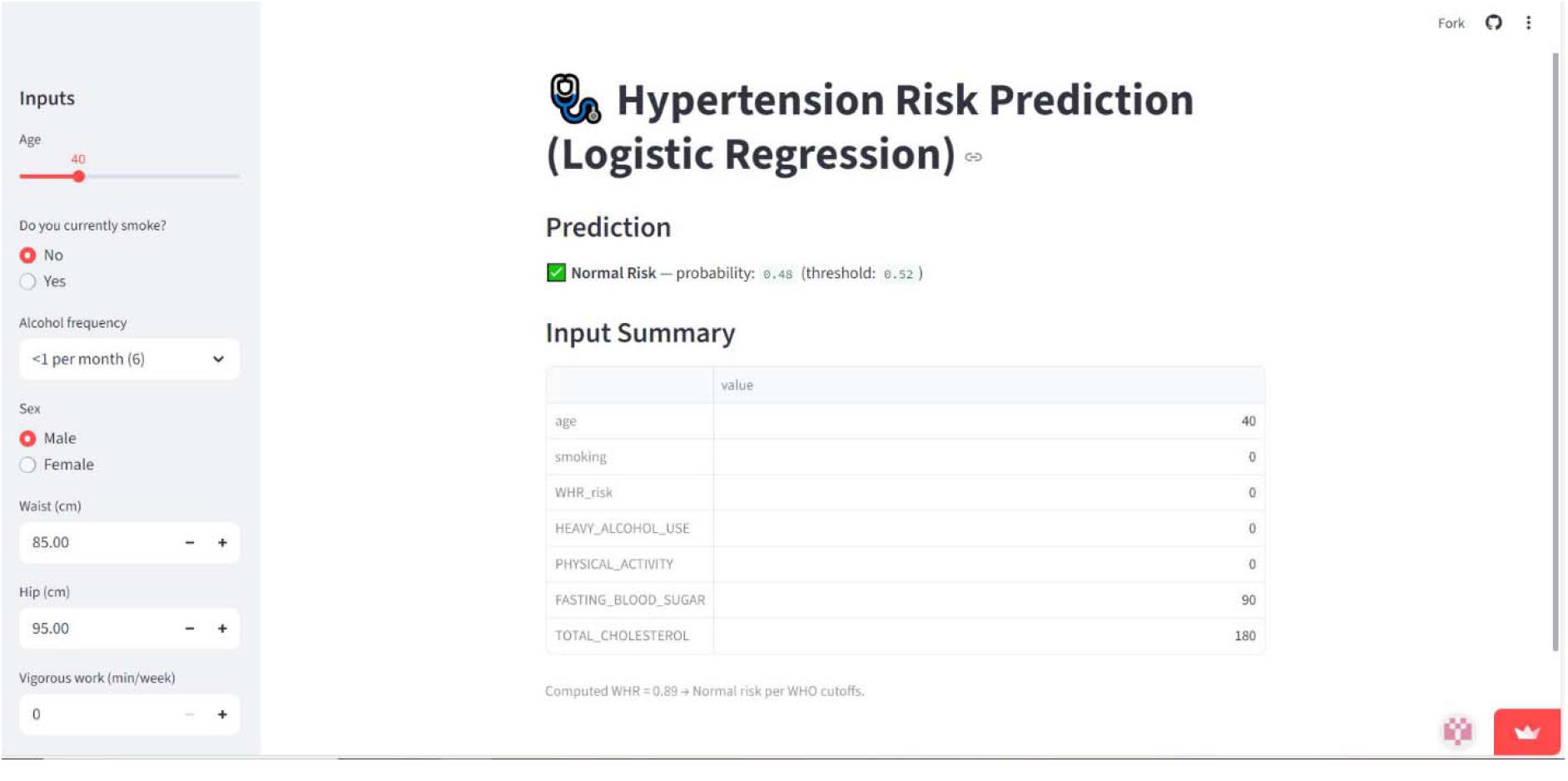
User friendly web application interface of Hypertension Risk Prediction.

## Discussion

### Principal Findings

This study developed and validated a hypertension risk prediction model using a rigorous machine learning pipeline and deployed it as a publicly available web tool. Our key finding is that Logistic Regression outperformed more complex algorithms, achieving the best discrimination (AUC ROC: 0.718), the best balance of precision and recall (F1-Score: 0.552), was well-calibrated, and demonstrated the highest clinical utility. The subsequent development of the Streamlit web application represents a critical translation of our research from a theoretical model into a practical tool ready for real-world screening and validation.

The seven-predictor model is parsimonious, actionable, and built on well-established risk factors. The use of SHAP values provides transparent, individualized explanations for predictions, a critical step towards building clinician trust. The deployment of the model via a web application directly addresses a common criticism of AI research in healthcare: the failure to move beyond journal publication into a format that can be tested and used in practice(9).

#### Comparison with Previous Work

Our finding that a linear model outperformed ensemble methods aligns with a growing body of literature suggesting that for structured tabular data with clear, linear risk relationships, simpler models can be superior due to their robustness, interpretability, and reduced risk of overfitting(10). In ethnic Chinese populations, clinical models incorporating gender, age, BMI, and blood pressure achieved AUCs of 0.732-0.741(11). Our emphasis on calibration, clinical utility (via DCA), and now implementation (via the web app) provides a more holistic view of model performance and impact that is often missing in ML health studies(12).

#### Strengths and Limitations

Our study has several strengths, including the use of a nationally representative sample, sophisticated handling of class imbalance, and threshold optimization. The creation of an interactive web application is a significant strength, enhancing the translational potential of our work and facilitating further validation through public use.

However, several limitations must be acknowledged. First, the model’s performance, while good, has a ceiling (AUC < 0.75). This is likely due to the absence of important predictors in the survey dataset, such as detailed family history, genetic markers, or longitudinal data on risk factors. Second, the model requires external validation in a prospectively collected, independent cohort to confirm its generalizability and true performance in a clinical workflow. Third, the cross-sectional nature of the survey data limits causal inference and may introduce bias from self-reported measures. Fourth, while the web app improves accessibility, its real-world impact depends on adoption rates, integration into existing health systems, and digital literacy of end-users. Finally, we relied on default hyperparameters for the ML models; a more extensive tuning process might have improved the performance of the tree-based models, though it is unlikely to surpass the well-calibrated and clinically useful Logistic Regression model for this specific task.

#### Implications for Practice and Research

The developed web application provides a immediate, no-cost tool for community health workers or primary care clinics to quickly screen individuals and prioritize them for confirmatory blood pressure measurement. This is particularly valuable in resource-limited settings where universal screening is challenging. By highlighting modifiable risk factors like WHR and physical activity, the tool can also serve as a platform for patient education and motivation for lifestyle changes.

Future research should focus on three key areas: 1) External validation: Prospectively validating the model and the web app’s performance in a new population and assessing its impact on clinical outcomes (e.g., rates of diagnosis, time to diagnosis). 2) Model refinement: Exploring the integration of additional variables to improve predictive power. 3) Implementation science: Studying the barriers and facilitators to the adoption of this tool in routine clinical and public health practice, particularly in low-resource environments.

## Conclusions

We successfully developed, validated, and deployed a hypertension risk prediction model as an accessible web application. By addressing key methodological challenges and focusing on implementation, we have demonstrated that a traditional Logistic Regression model can not only outperform more modern machine learning algorithms but also be transformed into a practical tool. The HTN Risk Nepal App bridges the gap between research and practice, offering a tangible solution for enhancing hypertension screening efforts and contributing to the global control of cardiovascular diseases. We encourage researchers and health professionals to utilize and validate this tool in their respective contexts.

## Data Availability

The dataset used is publicly available from WHO Data repository on request. The source code for the analysis and the web application is available at https://github.com/Dr-Charlie/web_app_deploy.git.

https://extranet.who.int/ncdsmicrodata/index.php/home

https://github.com/Dr-Charlie/web_app_deploy.git

## Declarations

### Ethics approval and consent to participate

Not applicable (publicly available, de-identified data).

### Consent for publication

Not applicable.

### Authors’ contributions

**Dr. Sandip Pandey**^**1**^: Conceptualization, Supervision, Project Administration, Writing – Review & Editing.

**Asmit Pandey**^**2**^: Methodology, Software, Formal Analysis, Investigation, Data Curation, Visualization, Writing – Original Draft Preparation.

**Dr. Aakash Neupane**^**1**^: Validation, Resources, Writing – Review & Editing.

**Dr. Deepak Subedi**^**3**^: Validation, Interpretation of results, Writing – Review & Editing.

**Dr. Aashish Guragain**^**1**^: Validation, Interpretation of clinical data, Writing – Review & Editing.

## Acknowledgements

Kritish Dhital, Everest Engineering college, Kathmandu, Nepal.

